# Assessing Pain Catastrophizing Through Free-Text Responses: A Validation of Large Language Models

**DOI:** 10.64898/2026.07.22.26358710

**Authors:** Angela Lee, Dokyoung Sophia You, Troy C. Dildine

**Affiliations:** Department of Anesthesiology, Perioperative and Pain Medicine, Stanford University School of Medicine, Stanford University, 1070 Arastradero Road, Suite 200, MC5596, Palo Alto, CA, 94304, USA; Health Promotion Research Center, Stephenson Cancer Center, and Department of Family and Community Medicine, University of Oklahoma, Oklahoma, United States

## Abstract

Validated measures of pain catastrophizing primarily assess catastrophizing as a stable trait. However, emerging evidence suggests catastrophizing fluctuates with context, highlighting a need for ecologically valid methods to capture it. This study evaluated large language models (LLMs) as implicit markers of catastrophizing from free-text responses from ninety-one adults with chronic pain receiving long-term opioid therapy (57.3% Female; mean age = 60.5 years). Patients completed baseline measures, including the trait pain catastrophizing scale (PCS), followed by a 10-minute writing task after random assignment to a negative, positive, or neutral pain-coping condition. State affect and pain were assessed before and after writing tasks and again after a cold pressor task (4ºC; ≤ 2 minutes). A state PCS followed the cold pressor task. Free-text responses were analyzed using four LLMs (Claude Opus 4; GPT Mini 4o; Llama 4 Maverick; and Gemini 2.5 Pro). ANOVA-based results supported discriminant validity, as all four LLM-derived pain catastrophizing scores differentiated negative from positive and neutral pain-coping conditions. Convergent validity was model-dependent; only Gemini-derived scores correlated with state catastrophizing (r = .22) and pain unpleasantness (r = .23). Divergent validity was mixed. LLM-derived scores were unrelated to pain intensity, but Gemini and Claude-derived scores showed small correlations with trait PCS (r’s = .21; 28, respectively). All LLM-derived scores also correlated with negative affect (r’s = .29–.41), comparable in magnitude to state PCS, suggesting limited specificity. These findings provide preliminary evidence that certain LLMs may serve as implicit markers of state pain catastrophizing, but further study is needed.

**Summary:** Leading Large Language Models (LLMs) demonstrate discriminant validity but mixed convergent and divergent validity scoring pain catastrophizing from persons living with chronic pain free-text responses.

## Introduction

Pain catastrophizing is a cognitive-affective response to pain characterized by persistent *rumination, magnification* of pain threats, and *helplessness* towards pain control [14]. Severe catastrophizing is associated with poorer responses pharmacologic and behavioral treatments for chronic pain and is linked to anxiety, depression, and greater pain severity and craving for opioid medication [19][12]. Pain catastrophizing has therefore gained attention as a key treatment target for improving outcomes in patients living with chronic pain [15].

Pain catastrophizing is commonly measured using validated quantitative instruments (e.g., Pain Catastrophizing Scale (PCS) [20]. The 13-item PCS was developed to measure catastrophizing as a relatively stable, trait-like pattern of thoughts toward pain. However, studies have shown that catastrophizing is influenced by situational context and may fluctuate within individuals over time, suggesting variability at the “state” level [18]. Consistent with this, within-person variability in pain catastrophizing tracks changes in pain outcomes [2]. To capture these momentary fluctuations, the 6-item state measure was developed. Nevertheless, this measure still relies on structured self-report and predefined items. In the present study, we instead examined patients’ free-text responses as a unique data source that may provide richer insight into current thoughts and personal pain experiences, serving as a potential implicit marker of state-based pain catastrophizing [13].

Analyses of unstructured text has evolved from manual scoring to lexicon-based sentiment analysis, traditional machine learning, and deep learning [25][10]. Sentiment analysis, among the most common computational tools for extracting subjective information from text, detects positive and negative affect content in language used across social media, community forums, and physician reviews [17]. However, sentiment analysis primarily quantifies broad emotional valence and does not capture specific cognitive patterns that characterize pain catastrophizing, such as pain-specific rumination, magnification, and helplessness. More recently, large language models (LLMs) have gained traction for analyzing free-text responses. Within chronic pain, classifiers trained on large datasets have improved pain diagnosis [16]. Beyond diagnosis, a recent proof-of-concept study in fibromyalgia demonstrated that LLMs can identify disorder-specific nuances in patients’ pain experiences from free-text responses[24]. Despite growing interest in LLMs [9], their application to pain catastrophizing remains largely unexplored. To our knowledge, no prior studies have examined LLMs as potential implicit markers for assessing pain catastrophizing from natural language. Further, most studies evaluate only one LLM, limiting model comparison.

In this study, we examined emerging LLMs to quantify free-text responses in patients with chronic pain on long-term opioid therapy who underwent experimental pain testing. Our primary aim was to validate the use of LLM-based methods for deriving pain catastrophizing scores by rigorously evaluating their correspondence with validated self-report measures of state pain catastrophizing. Secondary aims were to examine relationships with trait PCS, compare LLM approaches, and evaluate associations with other state measures, including negative affect and experimentally-induced pain. We hypothesized that LLM-derived catastrophizing scores would demonstrate discriminant validity (differing across conditions), convergent validity (associating with state pain catastrophizing and related constructs), and divergent validity (weak associations with pain intensity).

## Methods

### Participants

Participants were 98 adults (age ≥ 18 years) with chronic pain who were on long-term opioid therapy for at least three months prior to enrollment. Eligible individuals were required to have chronic pain lasting at least three months, be fluent in English, and indicate regular opioid use at a minimum frequency of once per week. Seven participants who did not write anything during the free-write portion were not scored. Therefore, the present study included 91 participants for the analysis. Exclusion criteria included active cancer, neurological disorders, suicidal ideation, medical conditions in the non-dominant hand (cold-pressor testing site), and having scheduled therapy appointments during the 2-week study observation period. Patients were recruited from the Pain Management Center at Stanford Health Care and from online advertisements. Initial eligibility was assessed via a structured phone interview, and informed consent was obtained from all eligible participants who elected to continue with the study. All lab appointments took place at the Stanford Systems Neuroscience and Pain Lab and The Institutional Review Board at Stanford University gave ethical approval for this work and all study procedures. Ninety-one participants attended and completed the lab visit. The mean age of participants was 60.5 years (± 11.9), and 57.3% of participants were female. The majority of participants were non-Hispanic White/Caucasian (81.3%), followed by Hispanic White/Caucasian (5.2%), African American/Black (5.2%), and Asian (5.2%).

### Experimental Design

This is a secondary analysis of an experiment conducted to examine the effect of pain catastrophizing on opioid craving in patients with chronic pain who were on long-term opioid therapy (clinicaltrials.gov: NCT04097743). The experiment design is summarized in Figure. 1. Participants were stratified by gender and craving for opioid medication (yes/no) and then, randomized to one of three expressive writing conditions: positive, negative, or neutral pain coping. After completing baseline assessments of trait and state measures, participants completed a 10-minute expressive writing task corresponding to their assigned condition, followed by post-writing state surveys. Participants then underwent the Cold Pressor Test (CPT, 4°C up to 2 minutes or as tolerated), after which they completed additional surveys administered immediately post-test and at 10-, 20-, and 30-minute follow-ups. Because this study aimed to validate the free-text analysis methods, we focus on survey data collected at baseline, immediately following the writing task, and immediately following the CPT.

**Figure 1.**
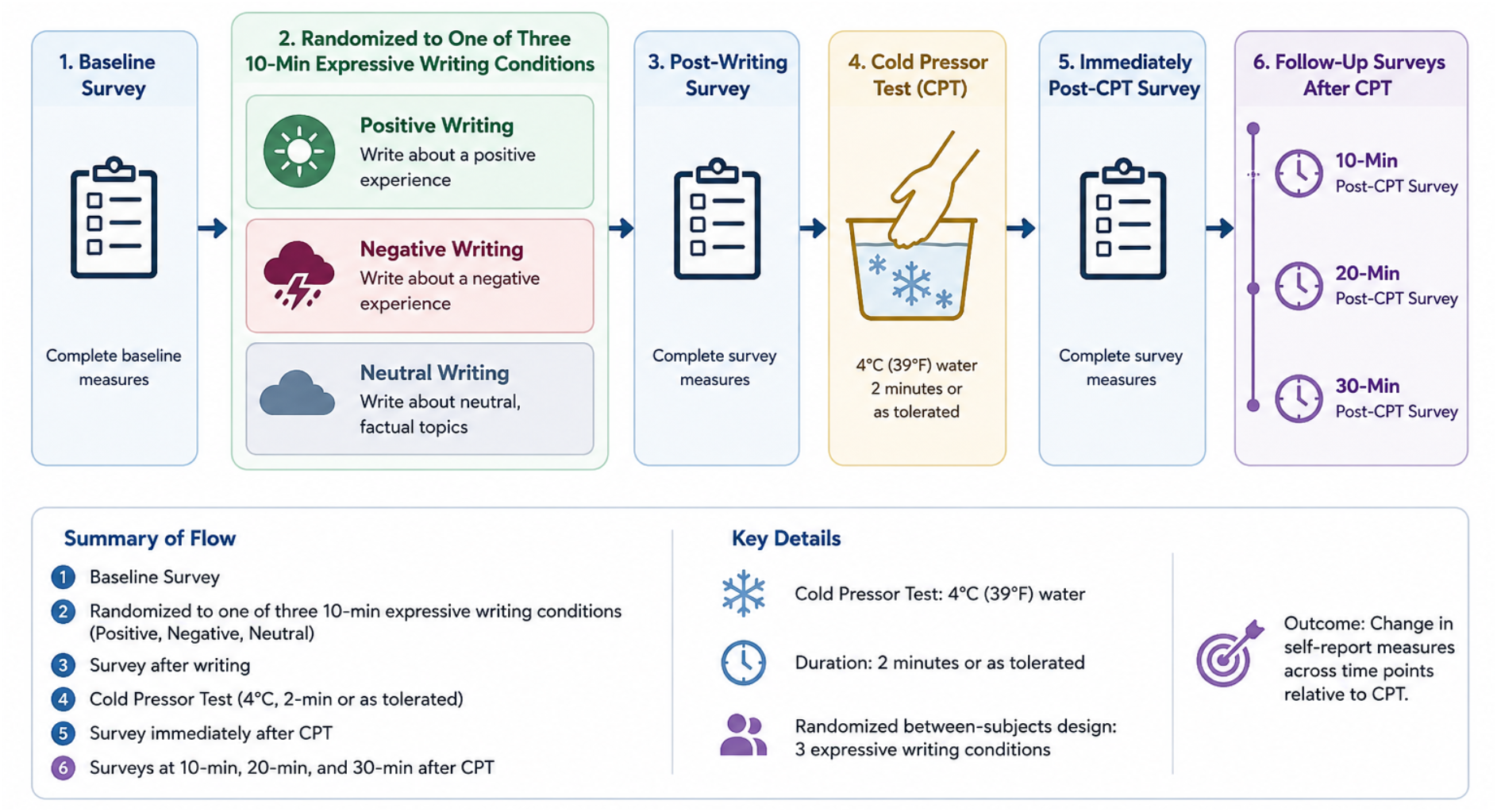
Experimental Design Overview (chatgpt generated image)

Although both pain catastrophizing and sentiment analyses were pre-registered: https://aspredicted.org/52wj4i.pdf, the present manuscript is limited to the evaluation of LLM-based pain catastrophizing scores because a) this study aimed to identify and quantify catastrophizing-relevant language in free-text responses and b) LLM-based pain catastrophizing scoring was considered a potentially more context-sensitive and construct-relevant approach than sentiment analysis for assessing pain catastrophizing as pressed in natural language. Analyses of LLM-based sentiment scores will be presented in a separate report.

### Free-text Writing Task

All participants were asked to write freely for 10 minutes in response to a prompt corresponding to the coping group to which they were assigned. Those assigned to the positive group responded to the prompt: “*Please think about a time when your pain was low. Then, please write down about your good pain day for 10 minutes (e*.*g*., *when was it, what did you enjoy doing, who were your supports, what made it a good day, what were helpful, any positive thoughts that came to your mind, how did you feel, what would you like to do if you have a good pain day again, etc*.).” Those assigned to the negative group responded to the prompt: “*Please think about a time you had the worst pain flare-up. Then, please write about your worst pain flare-up experience for 10 minutes (e*.*g*., *when was it, how bad it was, what were the triggers, what went wrong, what were the unknowns, were there any persons who were not helpful or not supportive, what made it worse, what negative thoughts about pain come to your mind, how did you feel, etc*.).” Those assigned to the neutral coping group responded to the prompt: “*Please write down what comes to your mind for the next 10 minutes*.*”* The experimenter left the room while the participant completed the writing task.

### Cold Pressor Test

Immediately after the writing task, participants in the positive and negative coping conditions selected a brief coping statement that corresponded to their assigned condition. Statements included sentiments such as “I can’t control my pain,” in the negative coping condition and “No matter how bad it gets, I can do this,” in the positive coping condition. Participants were also given the option to select their own positive or negative statement. Participants in the neutral condition received no instructions regarding coping statements. After choosing their statement, all participants underwent the cold pressor test (CPT), which involved immersing the non-dominant hand in 4°C water for up to two minutes or as long as tolerated [8]. Participants in the positive or negative coping conditions were instructed to repeat their chosen coping statement aloud throughout the CPT task. Participants could withdraw their hand at any time if the pain became intolerable.

### Measures

#### Pain Catastrophizing Scale (PCS)

The PCS [20] is a 13-item questionnaire used to measure trait-based catastrophizing at baseline. Patients were asked to indicate the frequency of catastrophizing-related thoughts from 0 – Not at all to 4 – All the time for each item. The total scores range from 0 to 52, with higher scores indicating greater pain catastrophizing (Cronbach alpha = 0.87, the 6-week test-retest *r* = 0.75) [23].

#### *Situational Pain Catastrophizing Questionnaire* (SPCS)

The sPCS is a 6-item measure assessing state pain catastrophizing in response to a specific situation [6]. In this study, it was administered immediately following the CPT. Each item is rated on a 0–4 scale, yielding total scores ranging from 0 to 24, with higher scores indicating greater pain catastrophizing in response to the CPT. The sPCS was used to assess whether the experimental manipulation effectively elicited condition-related differences in state catastrophizing following pain induction.

#### *The Spielberger State-Trait Anxiety Inventory* (STAI-S)

The 6-item short form of the Spielberger State Anxiety Scale, derived from the State–Trait Anxiety Inventory (STAI) [4][11]. It assesses current (state) anxiety (e.g., tension, worry, nervousness). Items are rated on a 4-point Likert scale, with three items reverse-coded. Total scores range from 0 to 12, with higher scores indicating greater state anxiety. The STAI-S was administered at baseline, post-writing, post-CPT, and at 10-, 20-, and 30-minute post-CPT follow-ups. However, STAI-S scores collected at baseline, post-writing, and post-CPT were used as an additional manipulation check to evaluate whether the experimental conditions influenced state anxiety.

#### *Self-Assessment Manikin* (SAM)

The SAM is a nonverbal pictorial measure of affect across three dimensions: valence, arousal, and dominance [1]. Each dimension is rated on a 9-point scale: valence (pleasant–unpleasant), arousal (calm–excited), and dominance (controlled–in control). Participants selected the figure that best represented their current emotional state. For this study, SAM responses collected at baseline, post-writing, and post-CPT were used to assess affective changes and manipulation effects across experimental conditions.

#### *Visual analogue scale* (VAS)

The VASs were used to assess current pain intensity and pain unpleasantness on a 0–100 scale. Pain intensity was rated from 0 (no pain) to 100 (most intense pain imaginable). Pain unpleasantness was rated from 0 (not at all unpleasant) to 100 (most unpleasant pain imaginable). For this study, VAS responses collected at baseline, post-writing, and post-CPT were used to evaluate condition-related effects. At baseline and post-writing, participants reported pain intensity and unpleasantness in relation to their own body pain. In contrast, post-CPT ratings reflected experimentally induced pain in the non-dominant hand following CPT.

#### LLM pain catastrophizing scores

Within Stanford Medicine’s (local) Secure AI Playground, text responses were scored with Anthropic’s Claude Opus 4; Open AI’s GPT Mini 4o; Meta’s Llama 4 Maverick; and Google’s Gemini 2.5 Pro. Texts were prefaced by the following prompt: “Please gauge and rate how much pain catastrophizing this text endorses from 0 - no catastrophizing to 1 - high catastrophizing. Pain catastrophizing is conceptualized as a negative cognitive–affective response to anticipated or actual pain. Please provide an overall score (i.e., 1 score value) for the entire quote and you can use decimals up to the hundredths place to provide specificity.” Initially, all texts were contained in one file and were scored at once. However, the results of batch-scoring the texts were inconsistent (e.g., empty text boxes received arbitrary values within the 0-1 range rather than receiving scores of 0 or NA). Due to this inconsistency, each text response was analyzed in unique prompt requests submitted each time by an experimenter. Further, without adding specificity on ability to provide decimals led to initial scores at 0 or 1. Each text response was analyzed in unique prompt requests, and the same prompt was used to analyze free-text responses using each of the LLMs. Stanford’s Medical Secure context is HIPPA compliant and allows for sensitive data to be handled. No other post-processing was applied to the LLM-based scores and the R scripts for all analyses and the LLM score outputs are available on osf upon request. Per [7], we adhere to all items for rigorous LLM application and reporting and note items B3-B5 (LLM configuration specifications like temperature and token usage, customization, and persistent memory) were not applicable or not available to assess in our local context and we report no competing interests with any of the included LLMs.

#### Validation Procedures and Analysis

As recommended [7], LLM-derived pain catastrophizing scores were validated against human ratings. First, three trained raters (AL, DY, and TD) evaluated participants’ free-text responses. Raters were blinded to participants’ experimental condition, and each narrative was independently evaluated by two raters. Pain catastrophizing was rated on a continuous scale from 0 (no evidence of pain catastrophizing) to 1 (extreme pain catastrophizing), based on the overall content of the narrative. Averages between the two human ratings served as the reference standard for evaluating the validity of LLM-derived pain catastrophizing scores.

Interrater reliability among human raters was assessed using intraclass correlation coefficients (ICCs). Convergent validity between human ratings and LLM-derived pain catastrophizing scores was evaluated using Spearman’s rank-order correlations. ICC values ≥0.70 were interpreted as indicating acceptable to good interrater reliability [21], whereas Spearman’s *r* values ≥0.30 were considered indicative of at least moderate convergent validity [22].

Discriminant validity was evaluated using one-way analysis of variance (ANOVA) to determine whether LLM-derived pain catastrophizing scores differed across the three experimental conditions. When the omnibus test was statistically significant, post hoc pairwise comparisons were conducted using Dunn’s tests to identify between-group differences.

Finally, convergent and divergent validity were examined by computing Spearman’s rank-order correlations between LLM-derived pain catastrophizing scores and validated self-report measures, including state and trait pain catastrophizing, state negative affect, and pain ratings. Convergent validity was supported by significant associations with theoretically related constructs, including state pain catastrophizing and state negative affect and anxiety. Divergent validity was evaluated by examining associations with theoretically less related constructs, including trait pain catastrophizing and pain ratings, with weaker or non-significant correlations expected. All scripts and LLM-scores are hosted on OSF and available upon request (link_upon_publication).

## Results

### Preliminary Validation of LLM-Derived Pain Catastrophizing Scores

Interrater reliability between the two human raters was excellent (ICC = 0.94, 95% CI [0.91, 0.96]), supporting the reliability of the human-coded benchmark used to evaluate LLM-derived pain catastrophizing scores. Correlations between human ratings and LLM-derived scores exceeded 0.30 (see Table 1), providing preliminary evidence of strong to very strong convergent validity between human and LLM-derived assessments of pain catastrophizing.

**Table 1.** Correlations between human-derived pain catastrophizing scores and LLM derived pain catastrophizing scores.

|  | *Correlation with human PCS* | *p-value* |
| --- | --- | --- |
| ***GPT Mini 4o*** | 0.86 | <.001 |
| ***Claude Opus 4*** | 0.80 | <.001 |
| ***Llama 4 Maverick*** | 0.85 | <.001 |
| ***Gemini 2.5 Pro*** | 0.73 | <.001 |

### Hypothesis 1 (Discriminant Validity): LLM-derived catastrophizing scores would differ across experimental conditions

A one-way ANOVA revealed a significant condition effect of writing condition on all four LLM-derived pain catastrophizing scores (Table 2). Post-hoc analyses indicated that LLM-derived pain catastrophizing scores were significantly higher in the negative coping condition than in both the positive (*p’s* < .05; see Figure 2 for a visual) and the neutral pain-coping conditions (*p’s* < .05). In contrast, LLM-derived scores did not differ significantly between positive and neutral coping conditions (*p’s* > .05). These results suggest that LLM-based pain catastrophizing scores demonstrate discriminant validity by distinguishing narratives reflecting negative pain coping from those reflecting positive or neutral pain coping. These results suggest that LLM-based pain catastrophizing scores demonstrate discriminant validity by distinguishing narratives reflecting negative pain coping from those reflecting positive or neutral coping. In contrast, neither trait PCS nor state PCS (SPCS) scores differed significantly across conditions (p’s > .05), suggesting that validated self-report measures were less sensitive to experimental condition effects than LLM-derived language-based scores.

**Table 2.**
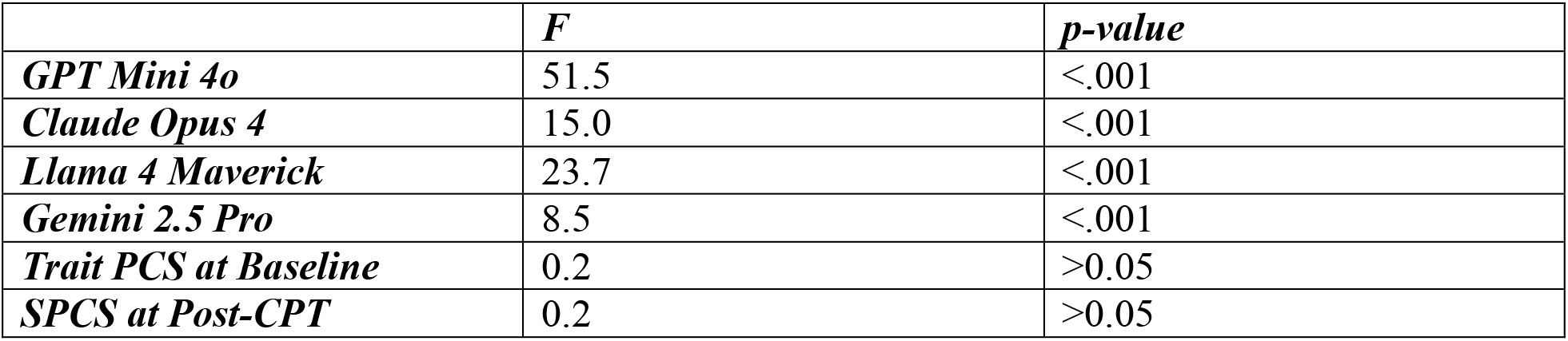
One-way ANOVA comparing pain catastrophizing among three conditions (n = 91)

**Figure 2.**
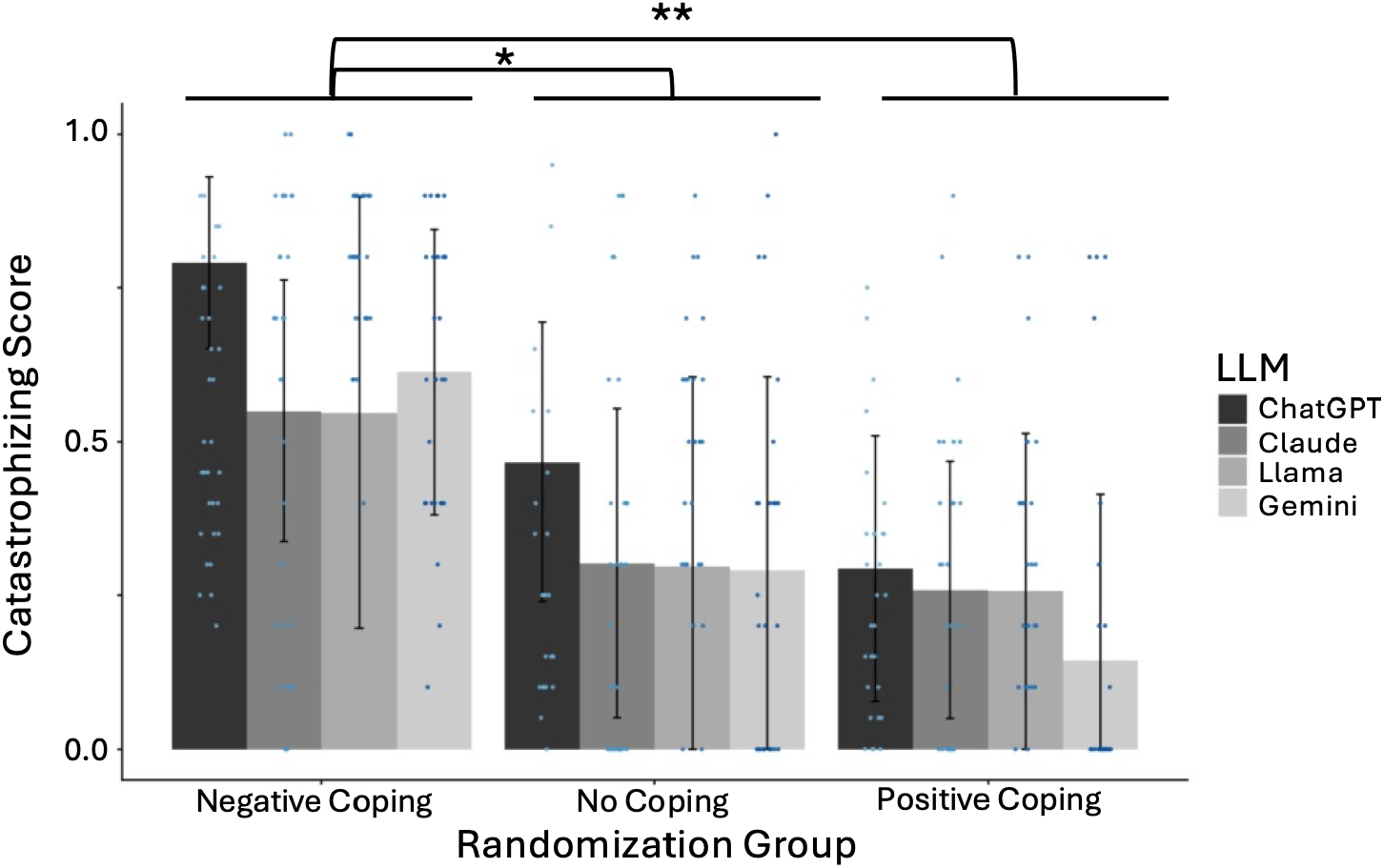
LLM-based pain catastrophizing scores by coping condition. Bar graphs of the average pain catastrophizing scores (y-axis) provided for each of the LLMs, grouped by coping condition (x-axis), are presented. Individual data points are also presented.

### Hypothesis 2 (Convergent validity): LLM-derived pain catastrophizing scores would show significant associations with state pain catastrophizing and general negative affect

Overall, findings provided weak support for convergent validity with associations with thematically relevant constructs of anxiety and negative affect, but little evidence for direct associations with our construct of interest, state-based pain catastrophizing.

### State Pain Catastrophizing

Surprisingly, only Gemini-based LLM-derived pain catastrophizing scores showed significant associations with state pain catastrophizing (r = .22), whereas the three other models all failed to meet significance (r’s < .19, see Figure 3 for confusion matrix).

**Figure 3.**
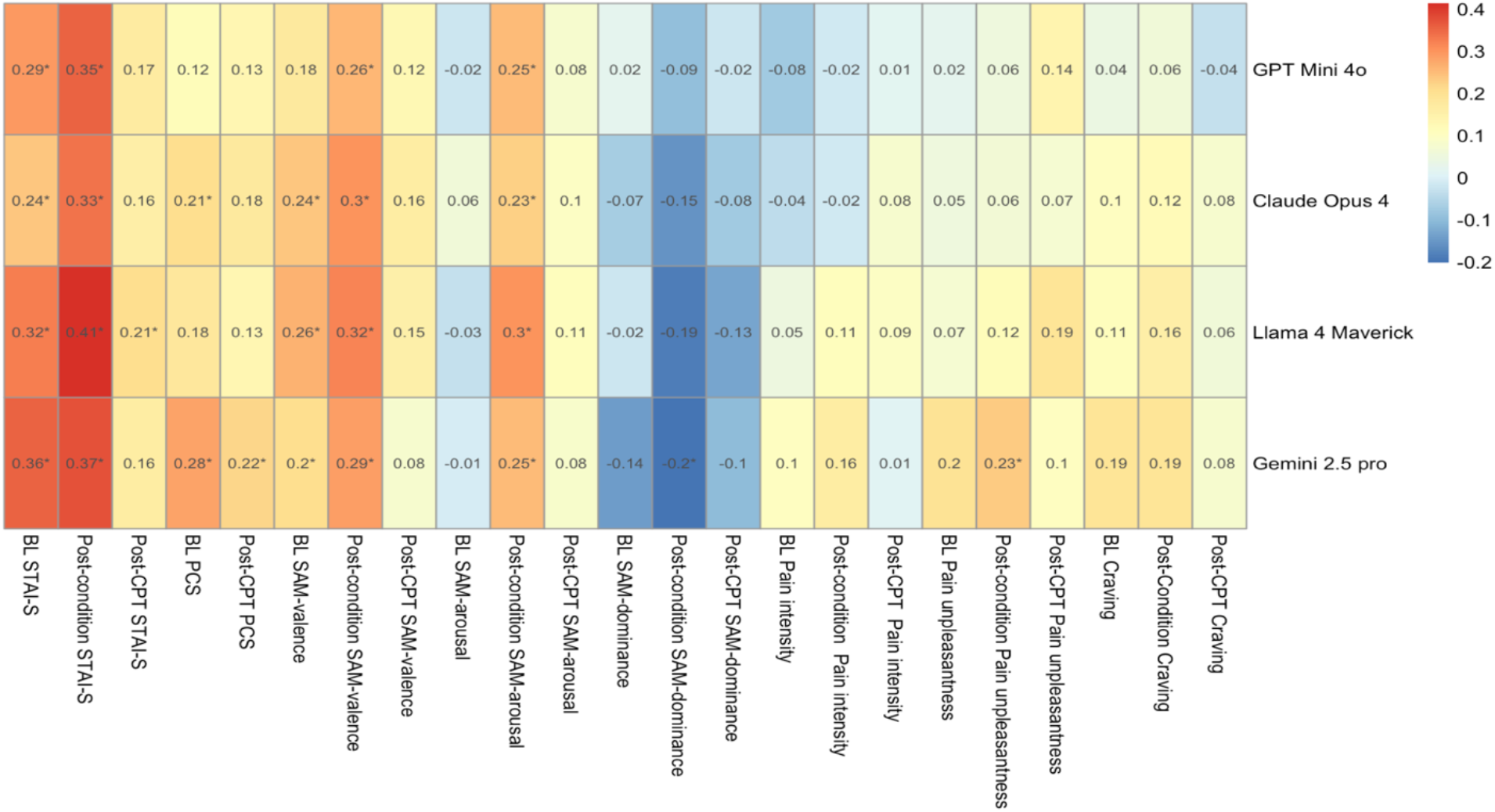
Heatmap of Correlation Coefficients Between LLM-Derived Pain Catastrophizing Scores and Validated State Measures. * p < .05. Each LLM-based PCS was correlated with each of our measures of anxiety (STAI), self-reported trait pain catastrophizing (BL-PCS) and state-based pain catastrophizing (Post-CPT PCS), valence, arousal, dominance, pain intensity, pain unpleasantness, and craving across different time points: baseline (BL), post-condition, and post cold pressor task (CPT).

### State anxiety (STAI-S)

LLM-derived scores showed consistent associations with state anxiety at baseline and immediately post-writing, with all four models showing significant correlations (r = .24–.41, p’s < .05). Following the cold pressor task, only Llama-derived scores remained significantly associated with state anxiety (r = .21, p < .05).

### Affective state (SAM)

Associations with affective state varied by dimension and time point. For valence, all LLM-derived scores except GPT were significantly associated with baseline ratings, and all four were significantly associated with post-writing valence. No significant associations emerged following the cold pressor task. For arousal, no baseline associations were observed; however, all four LLM-derived scores were significantly associated with post-writing arousal, with no associations following the cold pressor task. For dominance, no baseline associations emerged; following the cold pressor task, only Gemini-derived scores were significantly associated with dominance (r = −.20, p < .05).

### Hypothesis 3 (Divergent validity)

*LLM-derived pain catastrophizing scores would show weak or non-significant associations with trait catastrophizing and pain outcomes*. Overall, findings provided partial support for divergent validity, with a pattern of mixed and construct-dependent associations across measures and time points.

### Trait catastrophizing

Contrary to hypotheses, Gemini- and Claude-derived pain catastrophizing scores were significantly associated with trait PCS scores (r = .28 and .21, respectively, p’s < .05), whereas GPT- and Llama-derived scores were not. These associations, although small, suggest partial overlap with state-based pain catastrophizing. Notably, trait PCS and state PCS (SPCS) were significantly correlated in this sample (r = 0.37, p < 0.001), indicating expected convergence between trait and state self-report measures.

### Pain outcomes

LLM-derived pain catastrophizing scores showed minimal associations with pain outcomes. None of the models were significantly associated with pain intensity at any time point (all r’s < .17, see Figure 3). Only Gemini-derived scores were significantly associated with pain unpleasantness following the writing task (r = .23, p < .05).

## Discussion

Pain catastrophizing is increasingly understood to fluctuate with context, yet its measurement still relies almost entirely on the PCS, which does not capture how catastrophizing is expressed in patients’ own words. We evaluated whether four widely used large language models (LLMs) could quantify catastrophizing from brief free-text narratives in patients with chronic pain on long-term opioid therapy. The models agreed strongly with human raters, reliably detected experimentally manipulated coping context, and unexpectedly aligned more closely with trait than state catastrophizing. Findings support the limited and cautious use of language-based assessments for pain catastrophizing.

LLM scores matched human raters and detected context that self-report missed. All four models showed strong agreement with human judgments (r’s = 0.73 – 0.86), supporting their use as an efficient alternative to human coding. More strikingly, all four LLM-derived scores differentiated the negative coping condition from the positive and neutral conditions, whereas the state PCS self-report measure failed to distinguish among these conditions. However, these differences should be interpreted as reflecting measurement modality rather than superiority of one approach over another, as free-text analysis and self-report likely capture distinct aspects of the same underlying construct.

Despite this initial evidence supporting LLM scores as potential implicit markers for pain catastrophizing, evidence for convergent validity was limited and model-dependent. Only Gemini-derived scores showed significant associations with state pain catastrophizing, whereas other models showed minimal or no convergence. This suggests that model-specific characteristics may influence alignment with validated state measures. However, we note that there was presence of associations across all LLMs with other thematically similar constructs of negative affect and anxiety.

Associations with measures for divergent validity were also mixed. LLM scores were not associated with pain intensity, as predicted, and only Gemini scores associated with pain unpleasantness. However, associations with trait pain catastrophizing were observed with both Gemini- and Claude-derived scores (i.e., more than with state PCS). This suggests that a stable dispositional tendency toward catastrophizing emerges in language even when individuals are prompted toward opposing coping strategies. Written narratives may elicit reflective, meta-cognitive processing, whereas the state PCS likely captures immediate appraisal following acute pain; if so, the dissociation between LLM scores and state PCS is informative but raises questions about limitations in construct validity specificity for state-based PCS for chronic pain [5]. Initially, we hypothesized that state and trait PCS would be dissociable and appropriate for divergent validity, but we note that trait and state PCS were correlated (r = .37), consistent with theoretical overlap between state and trait pain catastrophizing.

The usefulness of LLMs in measuring pain catastrophizing faces current limitations. As described previously, the text entries must be entered in individual prompts to ensure accuracy. This approach is time-consuming and lacks scalability to larger datasets without specialized infrastructure and manual checks. Further, researchers using clinical and research data will need access to LLMs with verified security and HIPAA compliance, creating potential access barriers. Users with access to LLM APIs can typically access secure prompts and can automate the individual prompts; however, the cost of API usage can accumulate quickly and confidentiality and HIPAA compliance need to be verified. Beyond technical constraints, utility of LLMs may be limited due to their ‘black box’ nature (i.e., inability to articulate what defined the LLMs decision-making process [3]). This is particularly important to consider if interventions or clinical applications are an intended outcome.

Further exploration is needed to determine whether the usefulness of LLMs can be generalized to other types of responses, including unprompted texts or longer pieces of text. The current study assessed prompted writing in response to specific coping conditions, which may differ substantially from naturalistic patient responses. Studying clinical notes, patient portal messages and even social-media posts from individuals may provide more ecologically valid free-text data. Longitudinal applications are also needed, as we currently lack knowledge on whether LLM-based scores can predict future pain-outcomes or treatment adherence. Finally, there needs to be continual considerations made regarding the ethics of LLM and AI-based technology use in research and medical settings [26], with an understanding that algorithms contain biases and public-versions of models are not compliant and do not meet privacy and security thresholds necessary for clinical data.

### Conclusion

Our findings provide preliminary evidence that LLMs can capture state-level catastrophizing from brief free-text responses provided by patients with chronic pain. These models offer a novel and implicit approach that may complement traditional self-report measures by leveraging naturally occurring language expression. However, associations with trait pain catastrophizing suggest that LLM-derived scores may also reflect more stable or reflective cognitive tendencies and should not be assumed to index momentary pain catastrophizing exclusively. The state PCS should be reconsidered for its scope of construct validity and continued development, refinement, and validation of LLM-based approaches will be important for clarifying their construct and establishing their utility in pain psychology research and clinical applications.

## Data Availability

All scripts and LLM-scores are hosted on OSF and available upon request (link_upon_publication).

